# Risk factors for critical-ill events of patients with COVID-19 in Wuhan, China: a retrospective cohort study

**DOI:** 10.1101/2020.06.14.20130765

**Authors:** Sen Yang, Le Ma, Yu-Lan Wang, Qian Wang, Qiang Tong, Miao Chen, Hua Zhang, De-Hua Yu, Sheng-Ming Dai, Ran Cui

## Abstract

**Background:** Little is known about the risk factors for critical-ill events (intensive care, invasive ventilation, or death) in patients with COVID-19.

**Methods:** Patients with laboratory-confirmed COVID-19 admitted to the Wuhan Leishenshan Hospital from February 13 to March 14 was retrospectively analyzed. Demographic data, symptoms, laboratory values at baseline, comorbidities, treatments and clinical outcomes were extracted from electronic medical records and compared between patients with and without critical-ill events. The least absolute shrinkage and selection operator (LASSO) and multivariate logistic regression models were developed to explore the risk factors for critical-ill events. A risk nomogram was established to predict the probability for the critical-ill events. Survival analysis of patients with critical-ill events was performed by the Kaplan-Meier method.

**Results:** 463 COVID-19 patients were included in this study, of whom 397 were non-critically ill and 66 were critically ill (all from the intensive care unit). The LASSO regression identified four variables (hypersensitive cardiac troponin I, blood urea nitrogen, haemoglobin, and interleukin-6) contributing to the critical-ill events. Multivariable regression showed increasing odds of in-hospital critical-ill events associated with hypersensitive cTnI greater than 0.04 ng/mL (OR 20.98,95% CI 3.51-125.31), blood urea nitrogen greater than 7.6 mmol/L (OR 5.22, 95% CI 1.52-17.81, decreased haemoglobin (OR 1.06, 95% CI 1.04-1.10), and higher interleukin-6 (OR 1.05, 95% CI 1.02-1.08) on admission.

**Conclusions:** Hypersensitive cTnI greater than 0.04 ng/mL, blood urea nitrogen greater than 7.6 mmol/L, decreased haemoglobin, and high IL-6 were risk factors of critical-ill events in patients with COVID-19.

**Main point:** Hypersensitive cTnI greater than 0.04 ng/mL, BUN greater than 7.6 mmol/L, decreased haemoglobin, and high IL-6 were risk factors of critical-ill events (intensive care, invasive ventilation, or death) in patients with COVID-19.

## Introduction

An ongoing outbreak of the novel corona virus disease 2019 (COVID-19) pneumonia associated with the severe acute respiratory coronavirus 2 (SARS-CoV-2), started in December, 2019, in Wuhan, China, has spread rapidly around the world [1]. As of June 4, 2020, the total number of patients has risen sharply to 6.6 million around the world, with more than 380,000 deceased.

The clinical spectrum of COVID-19 pneumonia ranges from mild to critically ill cases. It is well established that patients with mild disease have fever along with respiratory signs and symptoms, such as dry cough [2]. Sepsis, respiratory failure, acute respiratory distress syndrome (ARDS), and septic shock are commonly observed in critically ill patients [3]. Epidemiological features, clinical presentation, and clinical outcomes of patients with COVID-19 have been well documented [4-7]. However, risk factors for critical-ill events (intensive care, invasive ventilation, or death) has not yet been well delineated. In this retrospective cohort study, we identified the risk factors associated with the critical-ill events among patients with confirmed SARS-CoV-2 pneumonia who were admitted to Wuhan Leishenshan Hospital. In addition, a prediction model of probability for the critical-ill events was established.

## Methods

### Study design and participants

This retrospective cohort study included two cohorts (non-critically ill and critically ill) of adult patients (aged ≥18 years) with COVID-19 from Wuhan Leishenshan Hospital. All patients who received intensive care or died or were discharged between February 13 and March 14, 2020, were included in this study. For patients who were alive by March 14, 2020, their living status was followed up and confirmed on March 28, 2020. The study was approved by the Research Ethics Commission of Zhongnan Hospital of Wuhan University.

### Data collection

Demographic, clinical, laboratory, treatment, and outcome data were extracted from electronic medical records using a standardised data collection form. All data were checked by two physicians (SY, LM) and a third researcher (RC) adjudicated any difference in interpretation between the two primary reviewers.

### Laboratory procedures

Methods for laboratory confirmation of SARS-CoV-2 infection have been described elsewhere [2]. Routine blood examinations were complete blood count, coagulation profile, serum biochemical tests (including renal and liver function, and electrolytes), myocardial enzymes, interleukin-6 (IL-6), C-reactive protein and procalcitonin.

### Definitions

Critically ill patients were defined as those admitted to the ICU who required mechanical ventilation or had a fraction of inspired oxygen (FiO_2_) of at least 60% or more [8]. Fever was defined as axillary temperature of at least 37.3°C. ARDS was diagnosed according to the Berlin Definition [9]. Sepsis and septic shock were defined according to the 2016 Third International Consensus Definition for Sepsis and Septic Shock [10]. Acute cardiac injury was diagnosed if serum level of high sensitivity cardiac troponin I (cTnI) exceeded upper reference limit set by Wuhan Leishenshan Hospital, or if new abnormalities were shown in electrocardiography and echocardiography. Acute kidney injury (AKI) was diagnosed according to the Kidney Disease: Improving Global Outcomes (KDIGO) clinical practice guidelines [11]. Acute liver injury was defined as elevated alanine aminotransferase (ALT) and/or aspartate aminotransferase (AST) and treatment was given determined by the treating physician. Hypoproteinaemia was defined as blood albumin of less than 40 g/L set by Wuhan Leishenshan Hospital. The disease severity status of COVID-19 was defined according to the Chinese management guideline for COVID-19 (version 7.0) [12].

### Statistical analysis

Continuous and categorical variables were presented as median (IQR) and n (%), respectively. Mann-Whitney U test, χ^2^ test, or Fisher’s exact test were utilized to compare differences between patients with and without critical-ill events where appropriate. To explore the risk factors associated with critical-ill events (intensive care, invasive ventilation, or death), LASSO and binary logistic regression models were used. The LASSO regression is a machine learning method that suitable for the reduction of high-dimensional data and collinearity. The LASSO regression model analysis was performed using the *glmnet* package of R. The receiver operating characteristic (ROC) with area under the curve (AUC) was used to determine accuracy of the LASSO. The prediction model that incorporated the independent risk factors was developed and presented as the nomogram. Nomogram was obtained using the *rms* and *regplot* packages of R. We used a Kaplan-Meier plot for survival data. Tests were two-sided with significance set at α less than 0.05. The Stata/SE 15.1 software (StataCorp, College Station, TX, USA) and R software 3.5.1 were applied for all analyses.

## Results

463 adult patients hospitalised in Wuhan Leishenshan Hospital with laboratory-confirmed COVID-19 were included in this study. 397 were non-critically ill and 66 were critically ill. 27 patients died during hospitalisation (all from the ICU) at 35 days. The median age of the 463 patients was 60.0 years (IQR 50.0-69.0) (Table 1). Hypertension was the most common comorbidity, followed by diabetes, and cardiovascular disease (Table 1). The most common symptoms on admission were dry cough and fatigue, followed by chest tightness, shortness of breath, myalgia, and fever (Table 1). Lymphocytopenia occurred in 36.1% (131/463) patients. Moderate to severe anemia occurred in 13.4 % (63/463) patients. The median time from illness onset to hospital admission was 10.0 days (IQR 5.0-17.0) and the median time from admission to death or discharge was 19.0 days (IQR 13.0-37.0) (Table 1).

**Table 1.** Demographic, clinical, and laboratory findings of patients with COVID-19 on admission.

|  | Total<br>(n=463) | Non-critically ill patients<br>(n=397) | Critically ill patients<br>(n=66) | p value |
| --- | --- | --- | --- | --- |
| **Demographics and clinical characteristics** |  |  |  |  |
| Age, years | 60.0 (50.0-69.0) | 58.0 (49.0-67.0) | 69.0 (60.5-79.5) | <0.0001 |
| Sex | .. | .. | .. | 0.007 |
| Men | 231 (49.9%) | 188 (47.4%) | 43 (65.2%) | .. |
| Women | 232 (50.1%) | 209 (52.6%) | 23 (34.8%) | .. |
| Any comorbidity | .. | .. | .. | .. |
| Diabetes | 80 (17.3%) | 62 (15.6%) | 18 (27.3%) | 0.02 |
| Hypertension | 179 (38.7%) | 146 (36.8%) | 33 (50.0%) | 0.041 |
| Cardiovascular disease | 34 (7.3%) | 20 (5.0%) | 14 (21.2%) | <0.0001 |
| Arrhythmia | 12 (2.6%) | 7 (1.8%) | 5 (7.6%) | 0.008 |
| Malignancy | 11 (2.4%) | 8 (2.0%) | 3 (4.5%) | 0.128 |
| Bronchial asthma | 7 (1.5%) | 6 (1.5%) | 1 (1.5%) | 0.584 |
| Cerebrovascular disease | 25 (5.4%) | 12 (3.0%) | 13 (19.7%) | <0.0001 |
| Chronic kidney disease | 18 (3.9%) | 11 (2.8%) | 7 (10.6%) | 0.002 |
| Chronic liver disease | 13 (2.8%) | 10 (2.5%) | 3 (4.5%) | 0.187 |
| Anemia* | 17 (3.7%) | 10 (2.5%) | 7 (10.6%) | 0.002 |
| CURB-65 score | 0.0 (0.0-1.0) | 0.0 (0.0-1.0) | 2.0 (1.0-2.0) | <0.0001 |
| Disease severity status | .. | .. | .. | .. |
| Mild | 3 (0.6%) | 3 (0.7%) | 0 | <0.0001 |
| General | 356 (76.9%) | 356 (89.7%) | 0 | .. |
| Severe | 38 (8.2%) | 38 (9.6%) | 0 | .. |
| Critical | 66 (14.3%) | 0 | 66 (100%) | .. |
| Deaths | 27 (5.8%) | 0 | 27 (40.9%) | <0.0001 |
| Time from illness onset to hospital admission, days | 10.0 (5.0-17.0) | 9.0 (4.0-15.0) | 11.0 (7.0-30.0) | 0.001 |
| Time from admission to death or discharge, days | 19.0 (13.0-27.0) | 20.0 (14.0-27.0) | 14.5 (7.3-21.8) | 0.005 |
| **Signs and symptoms** |  |  |  |  |
| Fever (temperature $\geq 37.3^{\circ}\text{C}$) | 51 (11.0%) | 32 (8.1%) | 19 (28.8%) | <0.0001 |
| Respiratory rate $>24$ breaths per min | 54 (11.7%) | 34 (8.6%) | 20 (30.3%) | <0.0001 |
| Pulse $\geq 120$ beats per min | 23 (5.0%) | 20 (5.0%) | 3 (4.5%) | 0.743 |
| Systolic blood pressure $< 90$ mm Hg | 0 | 0 | 0 | NA |
| Peripheral oxygen saturation, without inhaling oxygen ($\leq 93\%$) | 43 (9.3%) | 19 (4.8%) | 24 (36.4%) | <0.0001 |
| Dry cough | 316 (68.3%) | 281 (70.8%) | 35 (53.0%) | 0.004 |
| Sputum | 33 (7.1%) | 27 (6.8%) | 6 (9.1%) | 0.447 |
| Chest tightness | 101 (21.8%) | 83 (20.9%) | 18 (27.3%) | 0.246 |
| Shortness of breath | 117 (25.3%) | 100 (25.2%) | 17 (25.8%) | 0.922 |
| Dyspnea | 25 (5.4%) | 14 (3.5%) | 11 (16.7%) | <0.0001 |
| Myalgia | 68 (14.7%) | 64 (16.1%) | 4 (6.1%) | 0.033 |
| Fatigue | 162 (35.0%) | 143 (36.0%) | 19 (28.8%) | 0.254 |
| Anorexia | 35 (7.6%) | 32 (8.1%) | 3 (4.5%) | 0.452 |
| Diarrhoea | 34 (7.3%) | 31 (7.8%) | 3 (4.5%) | 0.451 |
| **Laboratory findings** |  |  |  |  |
| White blood cell count, $\times 10^9/L$ | 5.8 (4.6-7.1) | 5.6 (4.5-6.6) | 9.6 (6.6-12.6) | <0.0001 |
| <4 | 57 (12.3%) | 52 (13.1%) | 5 (7.6%) | <0.0001 |
| $\geq 4$ to <10 | 365 (78.8%) | 334 (84.1%) | 31 (47.0%) | .. |
| $\geq 10$ | 41 (8.9%) | 11 (2.8%) | 30 (45.4%) | .. |
| Lymphocyte count, $\times 10^9/L$ | 1.5 (1.1-1.9) | 1.6 (1.2-2.0) | 0.8 (0.5-1.3) | <0.0001 |
| <1.1 | 121 (26.1%) | 78 (19.6%) | 43 (65.2%) | <0.0001 |
| Haemoglobin, g/L | 122.0<br>(107.5-133.0) | 124.0 (113.0-135.0) | 85.0 (68.0-104.5) | <0.0001 |
| <60 | 9 (1.9%) | 1 (0.3%) | 8 (12.1%) | <0.0001 |
| $\geq 60$ to <90 | 53 (11.4%) | 20 (5.0%) | 33 (50.0%) | .. |
| $\geq 90$ | 401 (86.6%) | 376 (94.7%) | 25 (37.9%) | .. |
| Platelet count, $\times 10^9/L$ | 214.0<br>(172.8-270.0) | 220.0 (183.0-272.0) | 163.0 (78.0-260.0) | <0.0001 |
| <100 | 28 (6.0%) | 10 (2.5%) | 18 (27.3%) | <0.0001 |
| Alanine aminotransferase, IU/L | 23.0 (14.0-40.0) | 23.0 (14.5-39.0) | 22.5 (12.0-45.8) | 0.741 |
| >40 | 114 (24.6%) | 95 (23.9%) | 19 (28.8%) | 0.396 |
| Aspartate aminotransferase, IU/L | 21.0 (16.0-30.0) | 20.0 (16.0-27.0) | 31.0 (21.0-49.0) | <0.0001 |
| >40 | 69 (14.9%) | 46 (11.6%) | 23 (34.8%) | <0.0001 |
| Albumin, g/L | 37.9 (34.8-40.5) | 38.3 (35.7-40.8) | 32.2 (28.8-36.3) | <0.0001 |
| <40 | 335 (72.4%) | 275 (69.3%) | 60 (90.9%) | 0.0003 |
| Total bilirubin, $\mu\text{mol/L}$ | 9.4 (7.1-13.4) | 9.4 (7.2-13.0) | 9.1 (6.1-17.2) | 0.927 |
| >26 | 23 (5.0%) | 13 (3.3%) | 10 (15.2%) | 0.0005 |
| Creatinine, $\mu\text{mol/L}$ | 62.9 (52.4-75.8) | 62.1 (52.3-72.6) | 83.4 (55.0-199.6) | <0.0001 |
| >133 | 31 (6.7%) | 8 (2.0%) | 23 (34.8%) | <0.0001 |
| Blood urea nitrogen, mmol/L | 4.9 (3.8-6.3) | 4.7 (3.7-5.7) | 11.5 (5.8-19.7) | <0.0001 |
| >7.6 | 67 (14.5%) | 24 (6.0%) | 43 (65.2%) | <0.0001 |
| Creatine kinase, IU/L | 49.0 (34.0-74.3) | 48.0 (34.0-68.0) | 63.0 (34.3-143.8) | 0.005 |
| >185 | 20 (4.3%) | 11 (2.8%) | 9 (13.6%) | 0.001 |
| Creatine kinase-MB, ng/ml | 1.1 (0.8-1.8) | 1.0 (0.8-1.4) | 3.0 (2.2-6.2) | <0.0001 |
| >6.36 | 24 (5.2%) | 2 (0.5%) | 22 (33.3%) | <0.0001 |
| Lactate dehydrogenase, U/L | 190.0<br>(165.0-230.8) | 185.0 (162.0-211.0) | 329.0 (236.0-483.0) | <0.0001 |
| >245 | 108 (23.3%) | 63 (15.9%) | 45 (68.2%) | <0.0001 |
| Hypersensitive troponin I, ng/mL | 0.01 (0.01-0.01) | 0.01 (0.01-0.01) | 0.04 (0.02-0.24) | <0.0001 |
| >0.04 | 45 (9.7%) | 6 (1.5%) | 39 (59.1%) | <0.0001 |
| D-dimer, mg/L | 0.5 (0.2-1.26) | 0.4 (0.2-0.9) | 3.0 (1.4-7.4) | <0.0001 |
| <0.5 | 209 (45.1%) | 206 (51.9%) | 3 (4.5%) | <0.0001 |
| $\geq 0.5$ to <1 | 91 (19.7%) | 86 (21.7%) | 5 (7.6%) | .. |
| $\geq 1$ | 163 (35.2%) | 105 (26.4%) | 58 (87.9%) | .. |
| Prothrombin time, s | 11.4 (10.9-12.0) | 11.3 (10.8-11.6) | 13.2 (12.3-15.2) | <0.0001 |
| >13 | 52 (11.2%) | 13 (3.3%) | 39 (59.1%) | <0.0001 |
| Activated partial thromboplastin time, s | 27.2 (24.1-32.0) | 26.4 (23.6-29.8) | 38.6 (32.5-45.9) | <0.0001 |
| >40 | 38 (8.2%) | 10 (2.5%) | 28 (42.4%) | <0.0001 |
| C-reactive protein, mg/L | 1.5 (0.5-8.0) | 1.0 (0.5-3.8) | 46.3 (8.5-98.8) | <0.0001 |
| <4 | 262 (56.6%) | 250 (63.0%) | 12 (18.2%) | <0.0001 |
| ≥4 to <10 | 110 (23.8%) | 106 (26.7%) | 4 (6.1%) | .. |
| ≥10 | 91 (19.7%) | 41 (10.3%) | 50 (75.8%) | .. |
| Procalcitonin, ng/mL | 0.04 (0.03-0.07) | 0.03 (0.03-0.05) | 0.38 (0.13-2.88) | <0.0001 |
| <0.1 | 313 (67.6%) | 300 (75.6%) | 13 (19.7%) | <0.0001 |
| ≥0.1 to <0.25 | 23 (5.0%) | 10 (2.5%) | 13 (19.7%) | .. |
| ≥0.25 to <0.5 | 12 (2.6%) | 5 (1.3%) | 7 (10.6%) | .. |
| ≥0.5 | 115 (24.8%) | 82 (20.7%) | 33 (50.0%) | .. |
| IL-6, pg/mL | 5.4 (1.6-5.4) | 5.4 (1.5-5.4) | 108.7 (36.4-411.8) | <0.0001 |

Data are median (IQR), n (%), or n/N (%). *p* values were calculated by Mann-Whitney U test, $\chi^2$ test, or Fisher's exact test, as appropriate. COVID-19=coronavirus disease 2019. IL-6=interleukin-6.
MB=muscle and brain type. NA=not applicable.
\*Anemia events were recorded according to patients' history of present illness.

Patients who were non-critically ill received much more antiviral treatment (83.9% vs 45.5%) and Traditional Chinese medicine (84.4% vs 9.1%) than critically ill patients, whereas the critically ill patients received much more antibiotics, systematic corticosteroids, human serum albumin, renal replacement therapy, and convalescent plasma transfusion than non-critically ill patients (Table 2). 36 critically ill patients in ICU required invasive mechanical ventilation, of whom 20 (55.6%) died. Extracorporeal membrane oxygenation (ECMO) was used in four critically ill patients and two survived. Among critically ill patients, respiratory failure was the most frequently observed life-threatening complication, followed by acute cardiac injury, acute kidney injury, ARDS, and septic shock (Table 2).

**Table 2.**
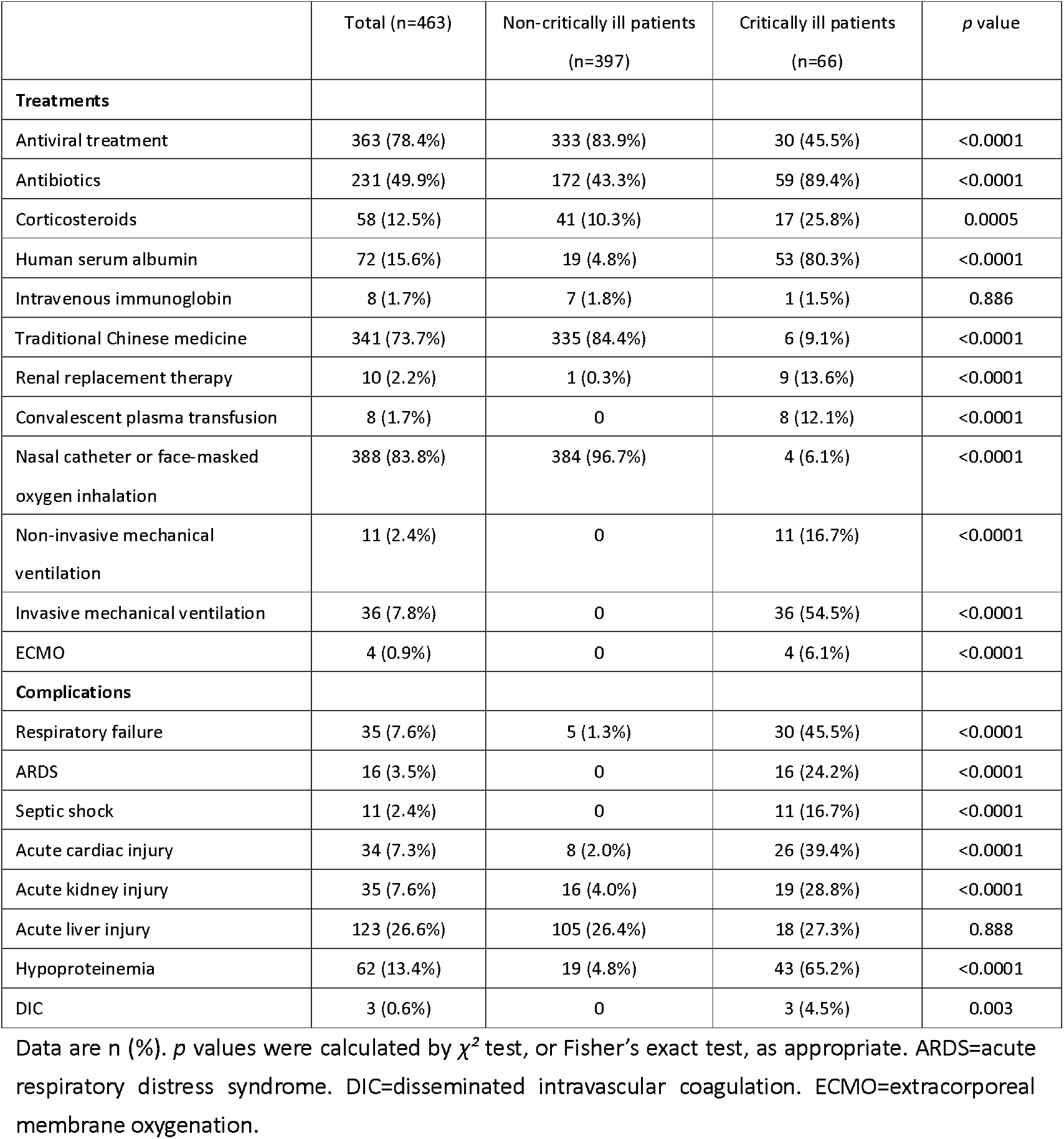
Treatments and complications.

Previous studies have shown male, older age, Sequential Organ Failure Assessment (SOFA) score, d-dimer, and the comorbidities such as hypertension, diabetes, and cardiovascular disease were associated with higher odds of critical events (intensive care or death)[3, 13-15]. In LASSO regression, we divided the full dataset (n=463, 66 with critical-ill events and 397 without) in a 7:3 ratio into training set (n=324) and validation set (n=139). A total of 19 variables (age, gender, comorbidities and laboratory data) were inputted and four variables (haemoglobin, cTnI, blood urea nitrogen (BUN), and IL-6) were finally selected in the training set (Figure 1).

**Figure 1:**
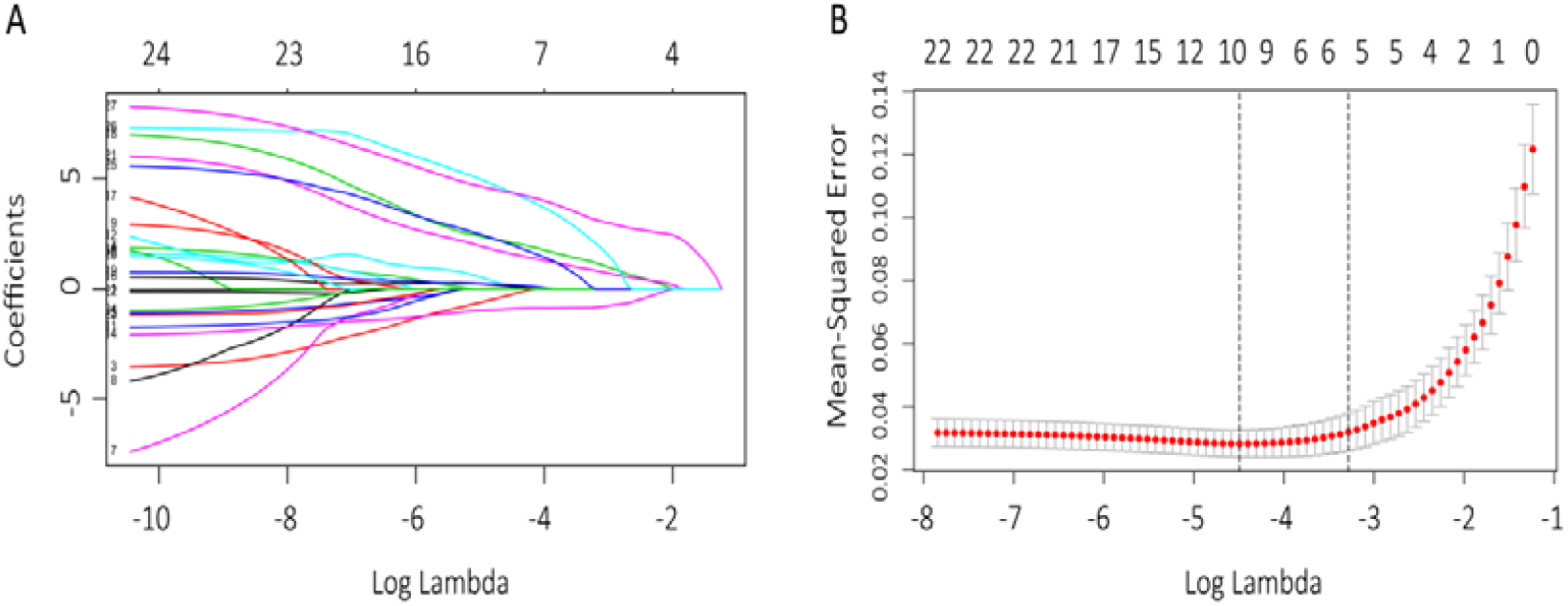
Clinical features selection via the LASSO regression model. The profile of coefficients in the model at varying levels of penalization coefficient (λ) plotted against the Log (λ) sequence, the top x-axis shows the average number of predictors (A). Identification of the optimal penalization coefficient (λ) in the LASSO model was performed via 10-fold cross-validation based on minimum criteria. The mean square error (MSE) was plotted verse Log (λ). Red dots represent average MSE for each model with a given λ, and vertical bars through the red dots show the upper and lower values of the MSE. The dotted vertical lines represent the optimal values of λ. When the optimal λ value of 0.034 with log(λ)=-3.38 was selected, the model reached the best. The upper and lower x-axes indicate the same meaning as in (A) (B). LASSO=least absolute shrinkage and selection operator.

The AUC in the training set was 0.984 (95% CI 0.959-1.000) and the AUC in the validation set was 0.959 (95% CI 0.898-1.000) (see Supplementary Figure 1), indicating that this feature selection model was of goodness of fit.

**Supplementary Figure 1:**
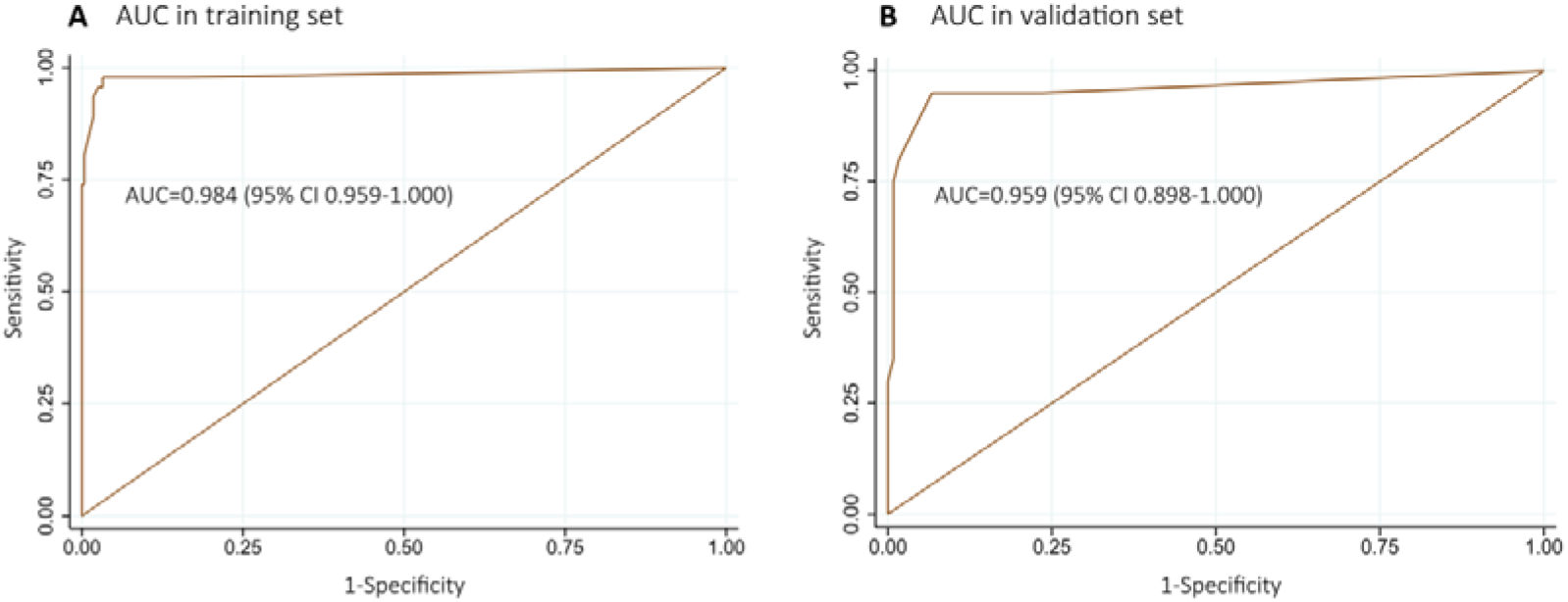
AUC to estimate the accuracy of the LASSO regression model. AUC in training set (n= 324) (A), and AUC in validation set (n=139) (B), both of two models showed high accuracy. AUC=area under curve. CI=confidence interval. LASSO=least absolute shrinkage and selection operator.

Based on the LASSO feature selection, the above four variables were included in the binary logistic regression, and we found that hypersensitive cTnI greater than 0.04 ng/mL (OR 20.98, 95% CI 3.51-135.31; *p*=0.001), BUN greater than 7.6 mmol/L (OR 5.22, 95% CI 1.52-17.81; *p=0.008*), decreased haemoglobin (OR 1.06, 95%CI 1.04-1.10, per 1 unit decrease (g/L); *p*<0.0001), and higher interleukin (IL)-6 (OR 1.05, 95%CI 1.02-1.08, per 1 unit increase (pg/ml); *p*= 0.001) at admission were significantly associated with increased risk of critical-ill events (Table 3). When adjusting for other confounders (CURB-65 score was excluded because itself included BUN), our model was validated as robust, because the four variables (haemoglobin, cTnI, BUN, and IL-6) were again selected as the risk factors contributing to the critical-ill events (see Supplementary Figure 2).

**Supplementary Figure 2:**
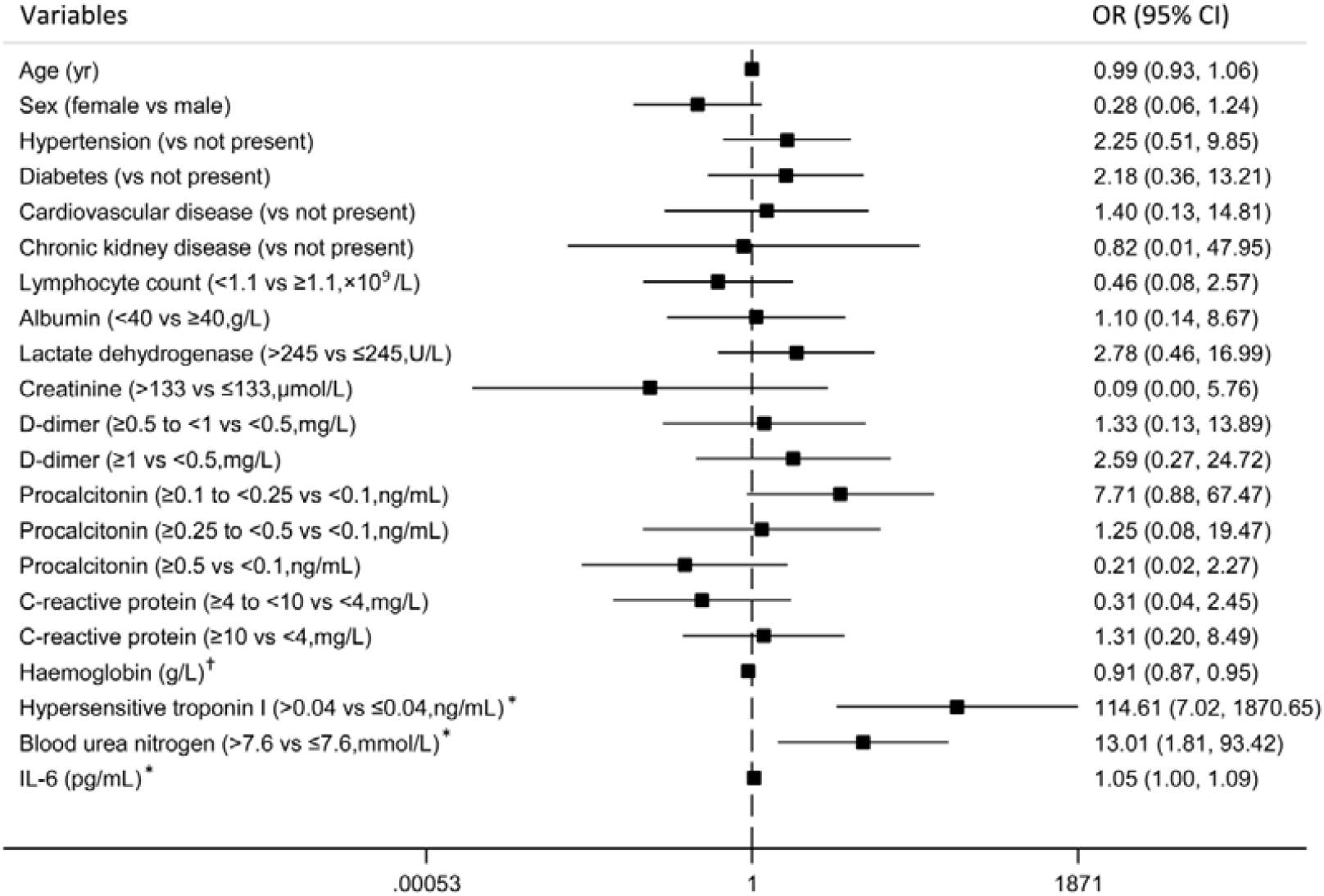
Re-validation of the binary regression by adjusting for other confounders. 17 variables were putted into the binary regression and four variables (haemoglobin, cTnI, blood urea nitrogen, and IL-6) were selected as the risk factors. CI=confidence interval. IL-6=interleukin-6. ^*^OR=odds ratio. *p*<0.05, ^†^ *p*<0.01.

**Table 3.** Risk factors associated with critical-ill events.

|  | Univariable<br>OR (95% CI) | *p* value | Multivariable<br>OR (95% CI) | *p* value |
| --- | --- | --- | --- | --- |
| **Demographics and clinical characteristics** |  |  |  |  |
| Age, years* | 1.06 (1.04- 1.09) | <0.0001 | .. | .. |
| Male vs female | 2.08 (1.21-3.58) | 0.008 | .. | .. |
| Comorbidity present (vs not present) |  |  |  |  |
| Diabetes | 1.67 (0.84- 3.30) | 0.142 | .. | .. |
| Hypertension | 1.72 (1.02-2.90) | 0.043 | .. | .. |
| Cardiovascular disease | 4.93 (2.22- 10.97) | <0.0001 | .. | .. |
| Arrhythmia | 3.33 (0.89- 12.54) | 0.075 | .. | .. |
| Malignancy | 2.08 (0.42- 10.28) | 0.371 | .. | .. |
| Bronchial asthma | 5.60 (2.23- 14.09) | 0.0003 | .. | .. |
| Cerebrovascular disease | 1.23 (0.13- 11.29) | 0.854 | .. | .. |
| Chronic kidney disease | 3.23 (1.05- 9.94) | 0.041 | .. | .. |
| Chronic liver disease | 1.43 (0.30- 6.92) | 0.653 | .. | .. |
| Anemia | 3.45 (1.07- 11.16) | 0.039 | .. | .. |
| **Signs and symptoms** |  |  |  |  |
| Respiratory rate, breaths per min |  |  |  |  |
| ≤24 | 1 (ref) | .. | .. | .. |
| >24 | 4.84 (2.57- 9.13) | <0.0001 | .. | .. |
| Peripheral oxygen saturation (without inhaling oxygen), % |  |  |  |  |
| >93 | 1 (ref) | .. | .. | .. |
| ≤ 93 | 6.06 (3.03- 12.12) | <0.0001 | .. | .. |
| CURB-65 score* | 4.92 (3.33- 7.26) | <0.0001 | .. | .. |
| **Laboratory findings** |  |  |  |  |
| White blood cell count, × 10<sup>9</sup>/L |  |  |  |  |
| <4 | 1 (ref) | .. | .. | .. |
| ≥4 to <10 | 0.97 (0.36- 2.59) | 0.944 | .. | .. |
| ≥10 | 28.36 (8.99- 89.46) | <0.0001 | .. | .. |
| Lymphocyte count, × 10<sup>9</sup>/L |  |  |  |  |
| ≥1.1 | 1 (ref) | .. | .. | .. |
| <1.1 | 7.65 (4.35- 13.43) | <0.0001 | .. | .. |
| Haemoglobin, g/L* | 0.93 (0.91- 0.94) | <0.0001 | 0.94 (0.91- 0.96) | <0.0001 |
| Platelet count, × 10<sup>9</sup>/L |  |  |  |  |
| ≥100 | 1 (ref) | .. | .. | .. |
| <100 | 14.51 (6.33- 33.25) | <0.0001 | .. | .. |
| Alanine aminotransferase, IU/L |  |  |  |  |
| ≤40 | 1 (ref) | .. | .. | .. |
| >40 | 1.29 (0.72- 2.30) | 0.397 | .. | .. |
| Albumin, g/L |  |  |  |  |
| >40 | 1 (ref) | .. | .. | .. |
| ≤40 | 4.44 (1.87-10.55) | 0.001 | .. | .. |
| Total bilirubin, μmol/L |  |  |  |  |
| ≤26 | 1 (ref) | .. | .. | .. |
| >26 | 5.28 (2.21-12.60) | 0.0002 | .. | .. |
| Creatinine, μmol/L |  |  |  |  |
| ≤133 | 1 (ref) | .. | .. | .. |
| >133 | 26.01 (10.96-61.71) | <0.0001 | .. | .. |
| Blood urea nitrogen, mmol/L |  |  |  |  |
| ≤7.6 | 1 (ref) | .. | .. | .. |
| >7.6 | 29.06 (15.12-55.84) | <0.0001 | 5.22<br>(1.53-17.81) | 0.008 |
| Creatine kinase, IU/L |  |  |  |  |
| ≤185 | 1 (ref) | .. | .. | .. |
| >185 | 5.54 (2.20-13.96) | <0.0001 | .. | .. |
| Lactate dehydrogenase, U/L |  |  |  |  |
| ≤245 | 1 (ref) | .. | .. | .. |
| >245 | 11.36 (6.37-20.37) | <0.0001 | .. | .. |
| Hypersensitive troponin I, ng/mL |  |  |  |  |
| ≤0.04 | 1 (ref) | .. | .. | .. |
| >0.04 | 94.13<br>(36.63-241.90) | <0.0001 | 20.98<br>(3.51-125.31) | 0.001 |
| D-dimer, mg/L |  |  |  |  |
| <0.5 | 1 (ref) | .. | .. | .. |
| ≥0.5 to <1 | 3.99 (0.93-17.08) | 0.062 | .. | .. |
| ≥1 | 37.93<br>(11.61-123.94) | <0.0001 | .. | .. |
| Prothrombin time, s |  |  |  |  |
| ≤13 | 1 (ref) | .. | .. | .. |
| >13 | 42.67 (20.38-89.35) | <0.0001 | .. | .. |
| C-reactive protein, mg/L |  |  |  |  |
| <4 | 1 (ref) | .. | .. | .. |
| ≥4 to <10 | 0.79 (0.25-2.49) | 0.683 | .. | .. |
| ≥10 | 25.41 (12.47-51.78) | <0.0001 | .. | .. |
| Procalcitonin, ng/mL* | 1.55 (1.21-1.97) | 0.0005 | .. | .. |
| IL-6, pg/ml* | 1.09 (1.07-1.12) | <0.0001 | 1.05 (1.02-1.08) | 0.001 |

CI=confidence interval. IL-6=interleukin-6. OR=odds ratio. \*Per 1 unit increase.

**Figure 2:**
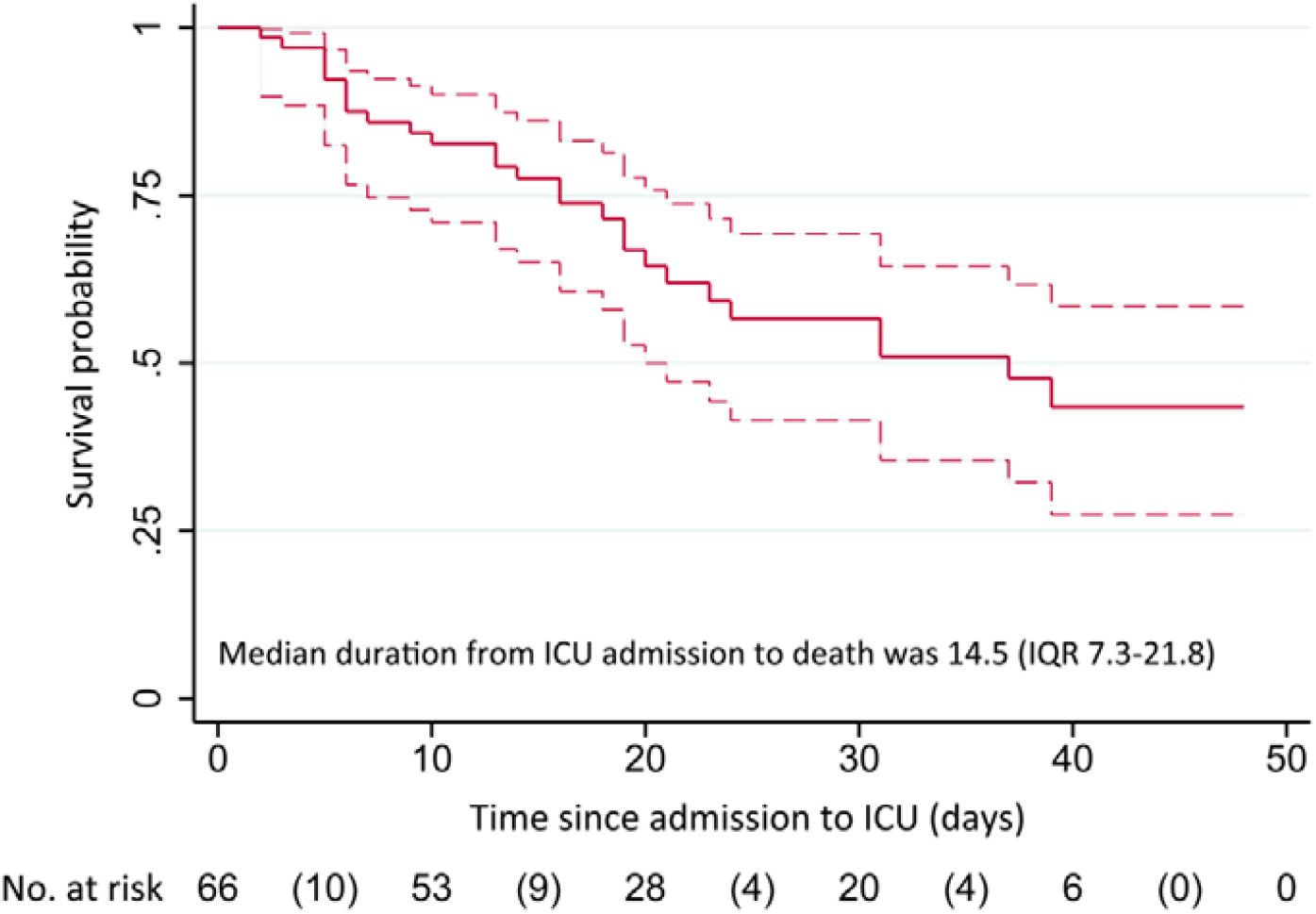
Survival probability of critically ill patients with COVID-19 in ICU. Dashed lines represent 95% confidence interval. Number in parentheses represent deaths. COVID-19=coronavirus disease 2019. ICU=intensive care unit.

Based on the selected risk factors, we established a risk model for evaluating the development of critical-ill events via nomogram (see Supplementary Figure 3). Calibration was employed with bootstrapping to decrease the bias of over-fitting in both training set and validation set. The internal-bootstrapped calibration plots for the probability of critical-ill events in training set and validation set showed an optimal agreement between prediction by nomogram (see Supplementary Figure 4).

For the death outcome, among 66 critically ill patients with COVID-19, 27 (40.9%) patients had died at 35 days, and the median duration from ICU admission to death was 14.5 (IQR 7.3-21.8) days (Figure 2). We further performed survival analyses stratified by cTnI, BUN, haemoglobin, and IL-6. High levels of cTnI, BUN, IL-6, and decreased level of haemoglobin were associated with poor survival probability (see Supplementary Figure 5).

**Supplementary Figure 3:**
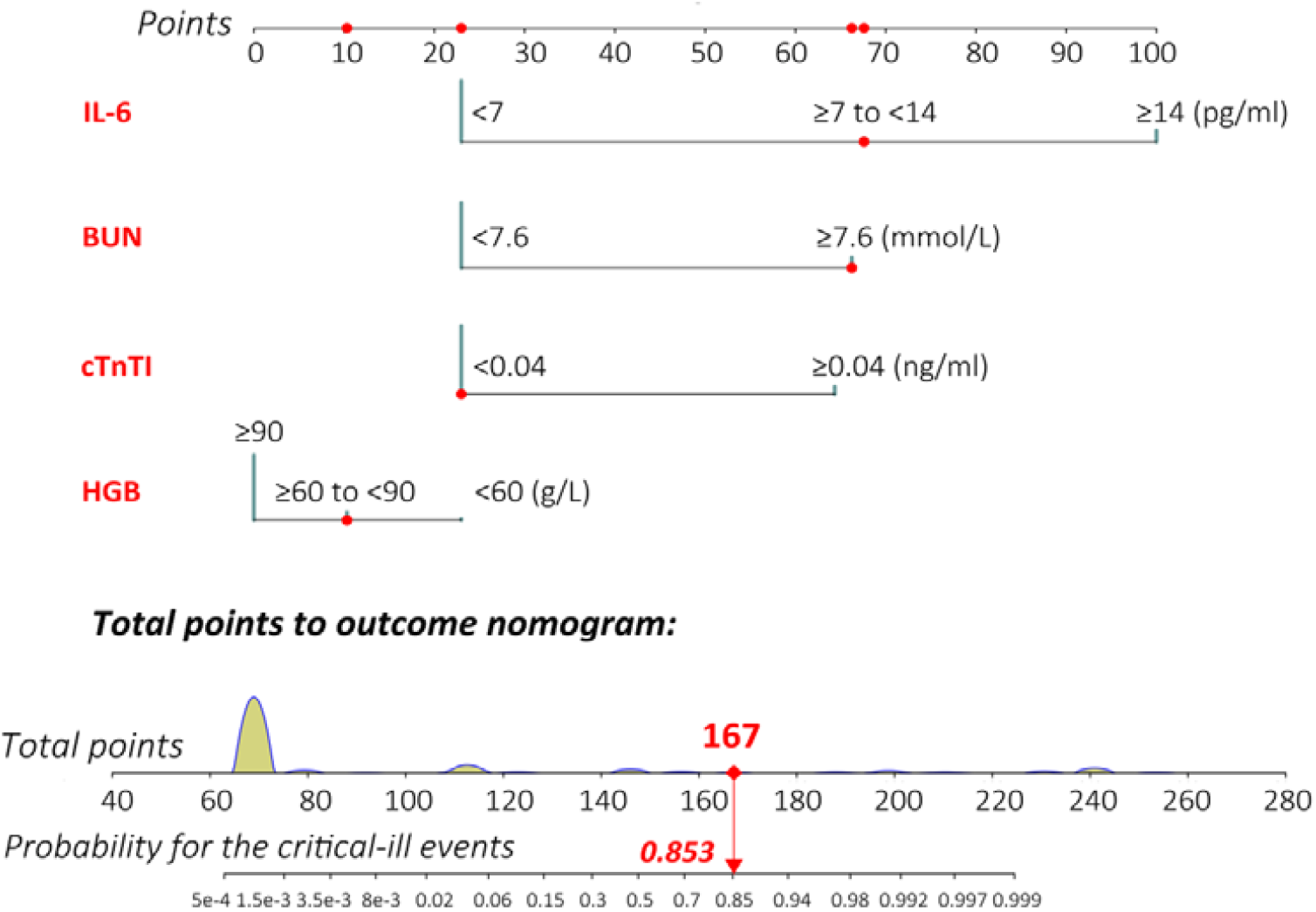
Nomogram to estimate the risk of critical-ill events in patients with COVID-19. Bars in blue display the distribution of patients in the training set (n=324). To calculate the total points of a specific patient, locate the value of each variable on the top point axis, add the points from all of the variables, and draw a vertical line from the total point axis to determine the probability at the lower line of the nomogram. Red track provided an example for the calculation of total-points-to-outcome. COVID-19=coronavirus disease 2019.

**Supplementary Figure 4:**
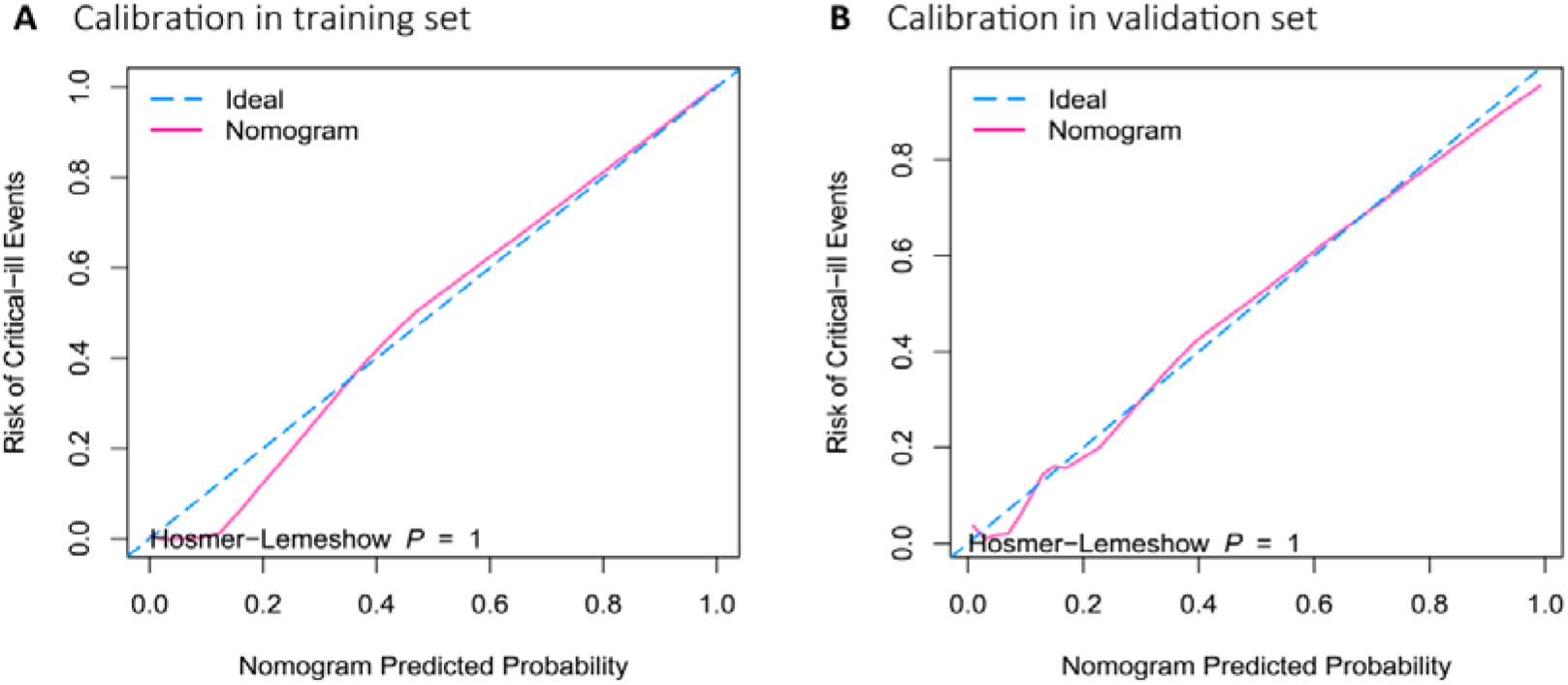
Calibration for the nomogram. Validity of the predictive performance of the nomogram in the training set (A) and validation set (B). Nomogram-predicted probability of critical-ill events is plotted on the x-axis and actual probability of critical-ill events is plotted on the y-axis. The 45-degree dashed line represented an ideal performance of the nomogram, in which the predicted outcome perfectly corresponded with the actual outcome. Both of validation in the training set and validation set showed high calibration (Hosmer-Lemeshow test *p*=1).

**Supplementary Figure 5:**
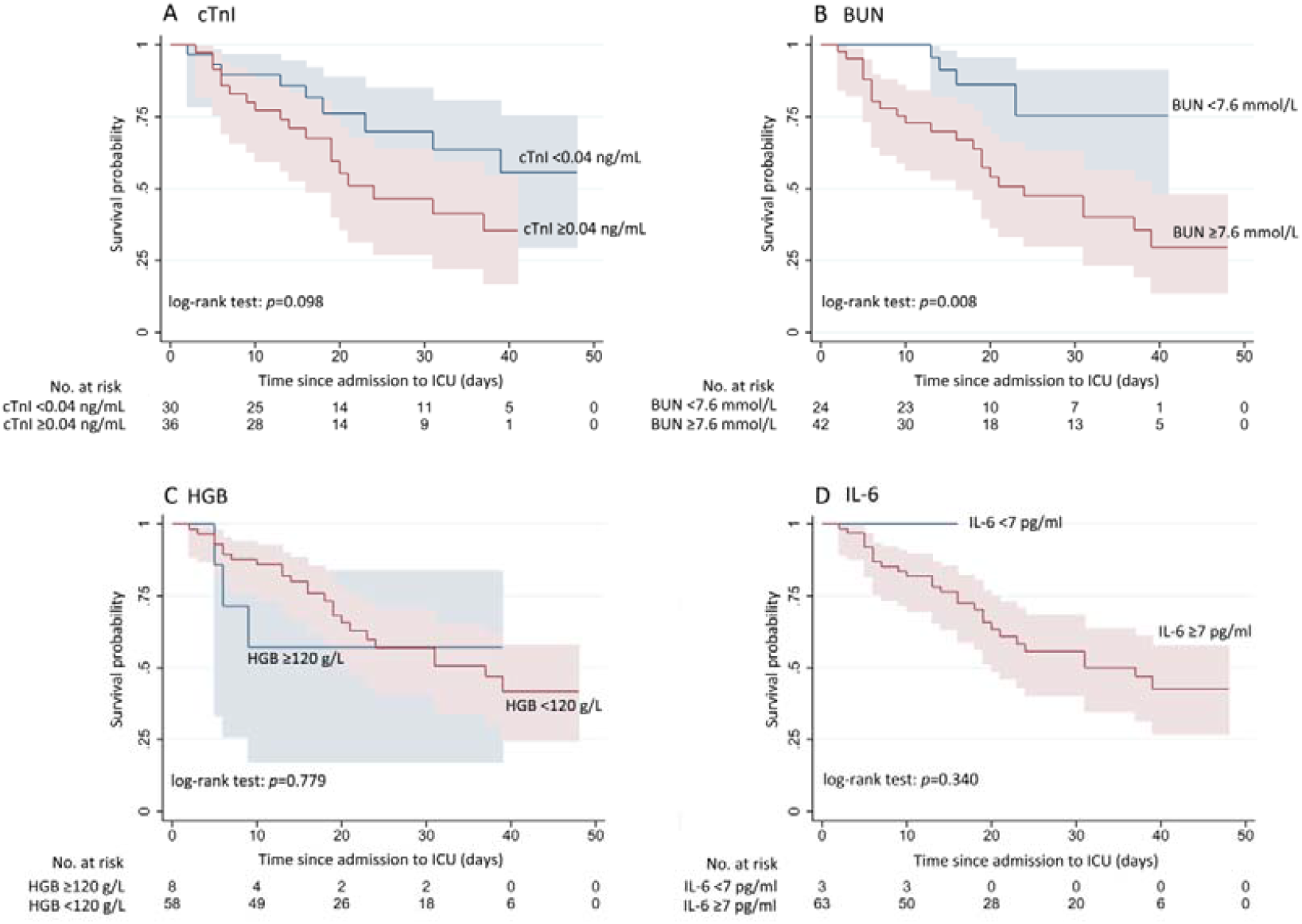
Survival analyses in 66 critically ill patients stratified by cTnI, BUN, haemoglobin and IL-6. High levels of cTnI, BUN, IL-6 and decreased level of haemoglobin were found to be associated with poor survival probability. BUN=blood urea nitrogen. cTnI=cardiac troponin I. HGB=haemoglobin. ICU=intensive care unit. IL-6=interleukin-6.

## Discussion

This retrospective cohort study identified four risk factors for critical-ill events (intensive care, invasive ventilation, or death) of patients with COVID-19 in Wuhan city. Hypersensitive cTnI greater than 0.04 ng/mL, BUN greater than 7.6 mmol/L, decreased haemoglobin, and higher interleukin IL-6 were found to be associated with increasing odds of in-hospital critical-ill events. Although the risk factors for mortality of adult inpatients with COVID-19 has been identified by a retrospective cohort study [3], the authors did not include some important factors (eg. IL-6 and procalcitonin) in multivariate logistic regression due to concern of the overfitting. In our study, these four risk factors were verified by LASSO and multivariate logistic regression models. Additionally, we developed and validated a prediction tool based on the selected four variables to evaluate the risk for critical-ill events for patients with COVID-19, which might aid in delivering timely treatment.

In COVID-19 patients with elevated inflammatory cytokines, postmortem pathology has revealed tissue necrosis and interstitial macrophage and monocyte infiltrations in the lung, heart and gastrointestinal mucosa [16]. In addition, exhausted lymphocytes with hyperactivated proinflammatory T cells [16] and decreased regulatory T cells [17] is commonly seen in critically ill patients, suggesting dysregulated immune responses. Among the excessive cytokines produced by activated macrophages, IL-6 is one of the key cytokines that closely related to the cytokine release syndrome (CRS), which may play a major role in the pathology of COVID-19. Elevated IL-6 levels were observed in patients with SARS coronavirus (SARS-CoV, a close counterpart of SARS-CoV-2) and were correlated with disease severity [18]. However, the elevated IL-6 levels were also observed in COVID-19 [2, 5, 19, 20] and might serve as a predictive biomarker for disease severity [21, 22]. In addition, maximal interleukin-6 levels (>80 pg/ml) before intubation showed the strongest association with the need of mechanical ventilation [23]. A prospective study regarding the tocilizumab (a monoclonal antibody that targets the interleukin 6 receptor) for the treatment of severe COVID-19 pneumonia with hyperinflammatory syndrome and acute respiratory failure showed significant clinical improvement [24]. In our study, IL-6 levels in critically ill patients were far more higher than that observed in non-critically ill patients. Extreme high IL-6 levels (>5,000 ng/mL) were found in two critically ill patients who were on ECMO and died due to CRS, coagulopathy, and respiratory failure.

Decline of hemoglobin was observed in 51% of 99 patients with SARS-CoV-2 infection reported by Wuhan Jin Yin-tan Hospital [5]. In the study of 1099 patients with COVID-19, the hemoglobin level of patients who reached composite endpoint (admission to ICU, requirement of invasive ventilation, and death) was lower than in those who did not (125.0 g/L (IQR 105.0-140.0) *vs* 134.0 g/L (120.0-148.0)) [25]. Inflammatory changes caused by SARS-CoV-2 infection might interfere with erythropoiesis, resulting in a decrease in hemoglobin. The low incidence of moderate or severe anemia in COVID-19 may attribute to the long life span of erythrocyte and the compensatory proliferation of erythrocyte induced by pneumonia associated hypoxia. In our study, among 66 critically ill patients, 41 (62.1%) had moderate to severe anemia (hemoglobin<90 g/L), 24 (36.4%) patients’ peripheral oxygen saturation was less than 93% without inhaling oxygen, and 36 (54.5%) patients received invasive mechanical ventilation. We speculated that, in critically ill circumstances, excessive inflammation could suppress the erythropoiesis, thereby hindering proliferation of erythrocyte induced by pneumonia associated hypoxia. Thus, the reduced hemoglobin levels might be an indicator of disease progression, and it would be more worthy of clinical attention.

AKI occurs frequently among patients with COVID-19. It occurs early and in temporal association with respiratory failure and is associated with a poor prognosis. Early reports from China found the rate of AKI in COVID-19 patients to range widely from 0.5-29% [2-6, 14]. Data from the New York hospital revealed that AKI developed in 36.6% (1,993/5,449) patients admitted with COVID-19 [26]. However, in critically ill or decreased patients, the incidence of AKI was much higher, up to 37.5-50% [3, 4]. In our study, 19 (28.8%) critically ill patients developed AKI and 9 (13.6%) patients received renal replacement therapy. BUN, the main component of azotemia caused by the AKI, has been listed as a risk factor associated with developing critical illness [27, 28]. Cox proportional hazard regression confirmed that elevated baseline BUN was an independent risk factor for in-hospital death (hazard ratio [HR] 3.97, 95%CI 2.57-6.14) [29].

Previous evidence substantiates the presence of acute cardiac injury in patients with COVID-19, which mainly manifested as an increase in high sensitivity cTnI levels (>0.028 ng/mL) [2]. However, acute cardiac injury might result in worsening arrhythmia, myocardial infarction or even cardiac arrest. It is notable that a observational study on 138 patients hospitalized with COVID-19 found that 7.2% of patients developed acute cardiac injury, and patients who received care in the ICU were more likely to have cardiac injury (22.2%) than non-ICU patients [6]. Consistently, our study found 39.4% of critically ill patients with cardiac injury. A large cohort study showed that patients with cardiac injury suffered more complications (including ARDS, AKI, electrolyte disturbances, hypoproteinemia, and coagulation disorders) and had higher mortality than those without cardiac injury (HR 3.41, 95%CI 1.62-7.16) [30]. The mechanism of acute myocardial injury might be related to the SARS-CoV-2 direct attack and the CRS. It is well established that SARS-CoV-2 binds with high affinity to human angiotensin-converting enzyme 2 (ACE2) which is used as an entry receptor to invade target cells [31]. ACE2 is widely expressed in both lungs and cardiovascular system [32] and, therefore, ACE2-related signalling pathways might play a role in heart injury. Other proposed mechanisms of myocardial injury include a CRS triggered by an imbalanced response by type 1 and type 2 T helper cells [33], and hypoxaemia caused by COVID-19 pneumonia, resulting in damage to myocardial cells.

Our study has certain strengths. To our knowledge, this is by far the largest respective cohort of hospitalized patients with COVID-19 with a focus on risk factors for the critical-ill events. In order to increase the reliability of the results, the LASSO with training and validation settings was utilized. Moreover, we constructed a risk nomogram with good calibration, which might help clinicians with early identification of patients who will progress to critically ill COVID-19 and enable better centralized management and early treatment of severe disease. However, some limitations existed in our study. First, due to the retrospective nature of the study, not all laboratory tests (including serum ferritin, IL-8, tumor necrosis factor-α and erythrocyte sedimentation rate) were done in all patients, especially in non-critically ill patients. Thus, their roles might be underestimated in predicting critical-ill events. Second, lack of effective antiviral drugs and late transfer from other community hospitals might have contributed to the development of critical-ill events in some patients, which may bias our results. Third, regarding the survival analyses, the p values for cTnI, haemoglobin, and IL-6 did not reach the statistical significance might due to inadequate follow-up and small samples. Further prospective study with large sample is warranted.

In summary, our data suggested that hypersensitive cTnI greater than 0.04 ng/mL, BUN greater than 7.6 mmol/L, decreased haemoglobin, and high IL-6 were risk factors of critical-ill events in patients with COVID-19.

## Data Availability

All data referred to in the manuscript can be obtained by contacting the corresponding author with reasonable request.

## Acknowledgments

We thank all patients and their families involved in the study.

